# Genome-Wide Association Study of Sepsis Susceptibility in ICU Patients Identifies Arab Founder Loss-of-Function Variant in *UBE2L3*

**DOI:** 10.64898/2026.09.25.26363954

**Authors:** Mahboubeh R. Rostami, Ali Ait Hssain, Amal Robay, Edward J Schenck, Jason J. Mezey, Ronald G. Crystal, Juan Rodriguez-Flores

## Abstract

Sepsis causes approximately 11 million deaths globally each year. Whole-genome sequencing (WGS) of founder populations offers a unique opportunity for discovery of genes related to susceptibility of sepsis. We performed GWAS and EXWAS on WGS data for n=210 Arab ICU patients (n=68 sepsis cases, n=142 controls, other ICU-related disorders) and prioritized n=40 associated deleterious founder variants in n=31 genes (REGENIE Firth regression p < 0.05; CADD score > 20; enriched AAF vs AllofUs/gnomAD), including a novel start-loss variant in UBE2L3 (c.2T>G; p.Met1?; CADD = 25.6; SIFT = 0.001) in sepsis cases (3/68 *vs* 0/142 controls; Fisher’s exact P = 0.026). In All of Us (AoU), UBE2L3 missense burden was associated with bacterial infection (n=26,939 cases; SKAT-O P = 2.38×10⁻³; OR=23.10; 95% CI = [0.63–845.27]). LLM AI models fine-tuned for drug development scored UBE2L3 a 7.8 of 10 as a drug target for sepsis, consistent with mouse UBE2L3 KO showing increased IL-1Beta mediated inflammation in sepsis.

## Introduction

Sepsis is a life-threatening organ dysfunction caused by a dysregulated host response to infection, defined by the Sepsis-3 criteria as a Sequential Organ Failure Assessment (SOFA) score of 2 or more points in the setting of suspected or confirmed infection [1]. Globally, sepsis accounts for approximately 11 million deaths annually, representing a major cause of morbidity and mortality in intensive care units (ICUs) worldwide [2]. Despite advances in critical care management, sepsis mortality remains high, and the identification of patients at greatest risk remains challenging.

Genetic factors are increasingly recognized as important determinants of sepsis susceptibility and outcome. Twin studies and family-based analyses have demonstrated a heritable component to infectious disease mortality[3]. Genome-wide association studies (GWAS) have identified several loci associated with sepsis-related phenotypes, but most studies have been conducted in European populations using genotyping arrays, which are inherently biased toward common variants and miss rare, population-specific alleles that may have large effect sizes[4]. The GWAS Catalog currently reports 62 genes associated with sepsis-related traits, but the genetic architecture of sepsis susceptibility remains incompletely characterized [5].

WGS in founder and understudied populations offers a powerful approach to identify novel disease-associated variants that may be enriched in specific ancestral backgrounds but absent from global reference databases. Arab populations, which have experienced historical bottlenecks and consanguinity, harbor founder alleles that are often absent from large-scale databases such as gnomAD [6] and the AoU [7]. These population-specific variants may contribute disproportionately to disease risk in these communities.

In this study we present a GWAS of sepsis susceptibility using WGS in n=210 Arab ICU patients from Qatar[8]. We performed single-variant association testing, sex-stratified analyses, gene-based burden testing, pathway analysis and independent gene-level validation to identify and characterize novel genetic determinants of sepsis susceptibility. The analyses identified suggestive associations of a compelling Arab founder loss-of-function variant in UBE2L3 with convergent evidence from predicted functional impact, population specificity and independent gene-level replication. Generative AI fine-tuned for drug discovery [9] identified the gene as an attractive target for sepsis, and mouse KO models of UBE2L3 in macrophages reported decreased ubiquitination and degradation of IL-1Beta, leading to enhanced inflammatory response in a sepsis model [10].

## Methods

### Study Population

Recruitment, sample collection, sequencing, and variant calling has been described in a previous manuscript [8]. Briefly, Hamad Medical Corporation (HMC) ICU patients (n=210) were recruited (IRB 19-00037) to participate in the study, comprising 68 sepsis cases and 142 non-sepsis ICU. WGS was performed on an Illumina HiSeq 4000 instrument to produce paired-end 2×150bp reads targeting 30x depth per sample, aligned to the GRCh38 human reference genome using BWA [11], and variant calling using GATK HaplotypeCaller [12]. Biallelic variants on autosomes (chromosomes 1–22) with a minimum minor allele count (minMAC) of 3 or greater were retained for downstream analysis, yielding a total of 16.6 million variants for association testing.

### Quality Control

Quality control of top association signals was performed using samtools mpileup (v1.20) across all 210 BAM files. For each variant, allele frequency concordance between expected (VCF-derived) and observed (pileup-derived) allele frequencies was assessed, with differences (deltaAF) of less than 0.03 considered acceptable. Strand bias was evaluated by comparing forward and reverse strand read proportions. Additionally, read-level verification was performed using Integrative Genomics Viewer (IGV) for top variant signals to confirm the presence of alternate allele reads and the absence of sequencing artifacts.

### Phasing

Genotype phasing was performed using BEAGLE v5.1 [13] to resolve haplotypes across the entire cohort. The merged VCF file containing unphased genotypes (0/1 notation) was processed to produce a phased VCF file with haplotype-resolved genotypes (0|1 notation). The phased output was indexed using tabix for downstream analysis. Cross-batch phasing was performed to ensure consistency across sequencing batches.

### Variant Annotation

Variants were annotated using a multi-tool pipeline. rsIDs were assigned using dbSNP [14]. Functional effect prediction was performed using SnpEff (GRCh38.99) [15], which provided gene annotations, loss-of-function predictions (LOF) and nonsense-mediated decay predictions (NMD). Clinical significance annotations were added from ClinVar [16]. Deleteriousness scores were obtained from dbNSFP v5.3.1a, which provided SIFT scores [17] and predictions, PolyPhen-2 HDIV scores [18], REVEL scores[19], MutationTaster predictions [20], FATHMM scores [21], MetaSVM scores [22] and Combined Annotation Dependent Depletion (CADD) Phred-scaled scores [23]. Population allele frequencies were annotated from gnomAD v4.1 [6]. Variants were further evaluated for presence in the AoU Research database.

### GWAS Analysis

Genome-wide association testing was performed using REGENIE v3.2.9 [4] in a two-step framework. In Step 1, a ridge regression model was fitted using a leave-one-chromosome-out (LOCO) approach to estimate a polygenic background effect, which served as the null model. In Step 2, Firth logistic regression was applied to test each variant for association with sepsis case-control status, accounting for the case-control imbalance (68 cases/142 controls = 32% case fraction). Covariates included age, sex, BMI, SOFA score, APACHE II score, and ICU days. The analysis was restricted to biallelic autosomal variants with minMAC ≥3, yielding 16.6 million variants for testing. Genomic inflation was assessed using the genomic control factor (λ_GC). Sex-stratified analyses were conducted separately in males (n = 111) and females (n = 99).

### Gene-Based Burden Testing

Gene-based burden testing [24] was conducted across n=20,414 genes using five statistical tests: SKAT (variance-component test, optimal for mixed effect directions), SKAT-O (optimal combination of SKAT and burden), SKATO-ACAT, ACATV (ACAT-V; aggregated Cauchy; burden-type), and ACATO (omnibus; combines BURDEN and SKAT optimally). Four variant masks were applied: M1 (loss-of-function only: stop-gained, frameshift, splice ±1/2, start/stop-lost), M2 (LoF + damaging missense with CADD ≥20), M3 (LoF + all missense) and M4 (synonymous variants, serving as a negative control). Variants were further stratified into four allele frequency (AAF) bins: singletons, AF < 0.1%, AF < 1%, and AF < 5%. This design resulted in 80 tests per gene (5 tests × 4 masks × 4 AAF bins). The Bonferroni-corrected significance threshold was p<6.8×10⁻⁷. Sex-stratified burden analyses were also performed.

### Founder Variant Prioritization and Independent Validation

Candidate founder variants were prioritized based on: (1) nominal p value of GWAS is significant; (2) predicted deleterious functional impact (CADD ≥20); (3) absence from gnomAD and AoU; (4) biological relevance to host immune pathways; and (5) enrichment in the Qatari ICU cohort compared to reference databases. Among 316 variants with CADD ≥20 and p ≤0.05, 40 variants across 31 genes were not found in AoU and were considered potentially unique to the Qatari population.

Independent gene-level validation of UBE2L3 was performed using gene-based burden testing (SKAT-O) of missense variants in 26,939 bacterial infection cases from the All of Us Research Program. This approach was used because variant-level replication was not possible for the population-specific UBE2L3 start-loss allele.

### Regional Linkage Disequilibrium Analysis

Phased r² was computed using PLINK2 v2.0.0-a.7.1 (--r2-phased --ld-snp-list --max-alleles 2 --snps-only) applied to the BEAGLE v5.1-phased WGS dataset. The analysis was restricted to biallelic SNPs within chr22:21,400,000–21,750,000 (GRCh38) with minimum allele count ≥1. A total of 4,480 variants were present in the region after joint genotyping; 860 multiallelic or indel variants were excluded, leaving 3,620 biallelic SNPs for LD analysis. Gene coordinates are from Ensembl release 109 (GRCh38.p13). The regional association and LD figure was generated in Python 3.12 using matplotlib v3.8.

### Drug Target AI

The nominally associated and predicted deleterious founder variants was used to prompt three large language model (LLM) systems for drug target prioritization: txgemma-9b, a foundation model fine-tuned for pharmaceutical target attractiveness using Therapeutic Data Commons [25] (including DisGeNet [26] scores for gene-disease links), gemma4-e4b, a recently released generalist foundation model and gemma4-12b-unified, an updated generalist foundation model. Each model was provided the summary statistics and functional annotation for the variants and prompted to score each gene based on drug target attractiveness on a scale of 0 to 10. The scores were compared across models, as well as within each model the prior (no association data) and post (with association data) scores were compared.

## Results

### Cohort Characteristics

The study cohort comprised 210 ICU patients, including 68 sepsis cases (32.4%) and 142 controls (67.6%). The cohort included 111 males (52.9%) and 99 females (47.1%), with ages ranging from 23 to 91 years (61.4 ±15.8) (Table 1). All three UBE2L3 p.Met1? carriers were male sepsis cases with organ dysfunction (SOFA ≥2).

**Table 1.** Cohort Characteristics^1^.

| **Characteristic** | **All (n=210)** | **Cases (n=68)** | **Controls (n=142)** |
| --- | --- | --- | --- |
| Sex (M/F) | 111/99 | 40/28 | 71/71 |
| Age, range (mean±sd) | 20 – 91 (59.2 ± 17.2) | 23 – 91 (61.4± 15.8) | 20 – 90 (58.2 ±17.7) |
| BMI range (mean±sd) | 4.1– 63.7 (30.6 ± 8.8) | 17.4 – 63.7 (30.1± 8.4) | 4.1 – 62.2 (30.9 ± 9.1) |
| SOFA score range (mean±sd) | 0 – 17 (7.7 ± 3.7) | 2 – 17 (9.0 ± 3.5) | 0 – 16 (7.1 ± 3.6) |
| APACHE II range (mean±sd) | 0 – 40 (20.9 ± 7.8) | 7 – 40 (22.4 ± 7.1) | 0 – 40 (20.2 ±8.0) |
| ICU days (mean±sd) | 1 – 406 (19.7 ± 33.4) | 2 – 124 (21.9 ± 23.2) | 1 – 406 (18.6 ± 37.3) |

<sup>1</sup> Shown is a description of the discovery cohort in this study, comparing all, cases, and controls. From top-to-bottom is sex, age range (in years), body mass index (BMI) range (m2/kg, Sequential Organ Failure Assessment (SOFA) score, APACHE II range, intensive care unit (ICU) days.

### GWAS Single Variant Tests

A total of 16.6 million biallelic autosomal variants with minor allele count (MAC) ≥3 were retained for association testing after GATK joint genotyping and VQSR filtering using REGENIE [4]. The genomic inflation factor was λ∼GC∼=0.840, indicating slight deflation consistent with the modest sample size (n=210) and conservative Firth logistic regression correction applied by REGENI (Supplemental Figure 1). No variants reached the conventional genome-wide significance threshold (p<5×10⁻⁸) (Figure 1), which was expected given the modest sample size of n=210 individuals. Four variants reached suggestive p < 5×10⁻⁵; Figure 1), and n=396,117 variants had nominal p values < 0.05. The n=4 suggestive association were common non-coding variants in genes and loci not previously associated with sepsis (STK32B, RP11-241F15.3, RP5-991O23.1, MUC19), suggesting these are artifacts given negative results in prior large sepsis GWAS studies of common variants [27–29]. There were n = 316 M1 (predicted loss of function or pLoF) or M2 (pLoF+pDM or predicted deleterious missense) variants association with n= 295 with p < 0.05 (Supplemental Table I, II).

**Figure 1.**
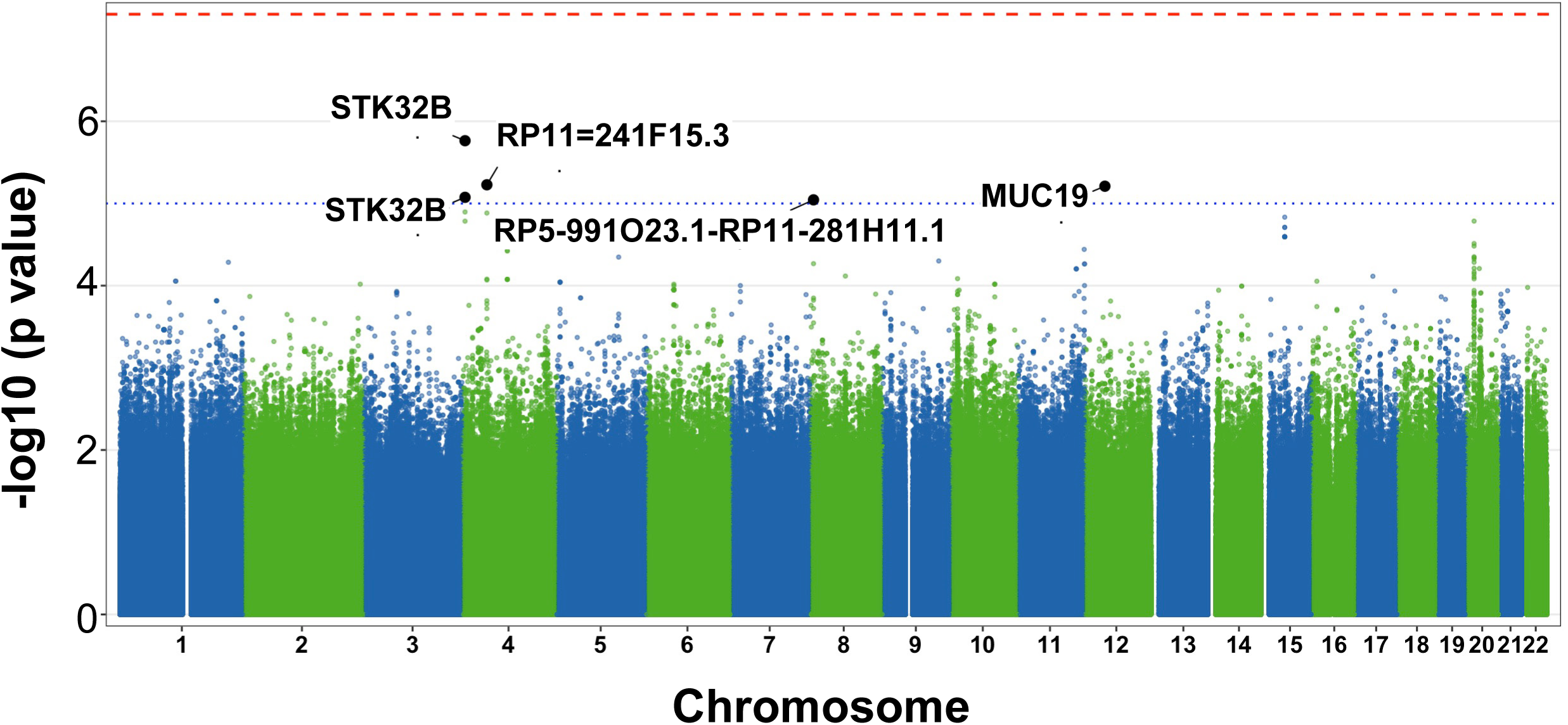
Manhattan plot of genome-wide association results. Single-variant association results for sepsis susceptibility in 210 ICU patients (68 cases, 142 controls). The x-axis shows chromosomal position and the y-axis shows -log10(p-value). The dashed red line indicates genome-wide significance (p < 5×10⁻⁸) and the dashed blue line indicates the suggestive threshold (p < 1×10⁻⁵). Top loci are labeled: *STK32B* (chr4p16.2), *RP11-241F15.3* (chr4q13), *MUC19* (chr12q12), and chr8p23.1 (*DEFB* cluster).

### Gene-Based Burden Tests

Gene-based burden testing pools association signal across rare variants. Across n = 20,414 genes there were no tests passing the Bonferroni multiple testing correction threshold with alpha = 0.05 (p < 2.4×10-6) (Figure 2). Several biologically relevant associations were found in this subset that reinforced the single variant association test results. The top novel signal was TRPM5 (transient receptor potential cation channel subfamily M member 5) under the M2 mask (LoF + damaging missense), with an OR of 7.43 (p = 3.4×10⁻⁴). TRPM5 showed a female-enriched effect (female OR = 10.6, p = 0.012). TRPM5 may act as a regulator of immune-cell activation and inflammatory signaling, suggesting a potential role in the dysregulated inflammatory response that characterizes sepsis [30]. IL19, an anti-inflammatory interleukin and IL-10 family member [31], showed association under the M3 mask (LoF + all missense; OR = 2.65, p = 7.1×10⁻⁴). GZMM (granzyme M), involved in natural killer cell cytotoxicity[32], was associated under the M3 mask (OR = 14.0, p = 8.7×10⁻⁴).

**Figure 2.**
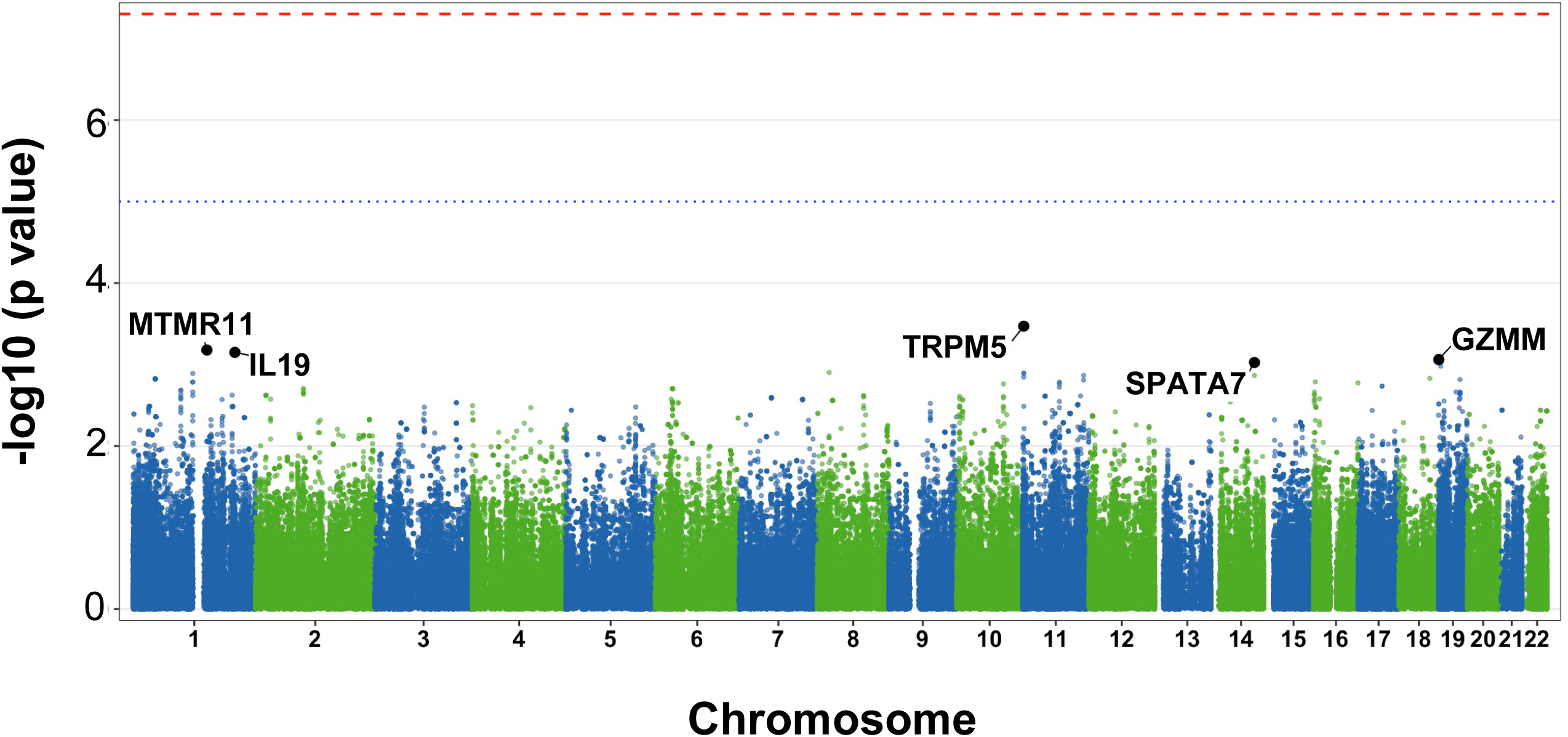
Manhattan plot for burden test. The x-axis shows chromosomal position and the y-axis shows -log10(p-value). The dashed red line indicates genome-wide significance (p < 5×10⁻⁸) and the dashed blue line indicates the suggestive threshold (p < 1×10⁻⁵). The five most significant genes are labeled: *MTMR11* (chr1), *IL19* (chr1), *TRPM5* (chr11), *SPATA7* (chr14), and *GZMM* (chr19).

### Identification of an Arab Founder Loss-of-Function Variant in *UBE2L3*

Deleterious founder alleles provide a promising route to target discovery in this small cohort of variants and genes not previously associated with sepsis in prior studies. Among n=316 variants with predicted deleteriousness (CADD ≥20) and nominal significance (p ≤0.05), n=40 variants predicted deleterious to n=31 genes were absent from AoU and GnomAD (Table 2, Supplemental Table III). The n=31 genes also with burden test p value < 0.05 included: *FAM72B, NOTCH2NL, SEC22B, BOD1, HLA-B, ZNF92, DYNLL1, GXYLT1, PABPC3, ZFYVE1, ZNF443*, and *UBE2L3* (Supplemental Table IV). Systematic evaluation of these variants for biological plausibility using publicly available large language models identified UBE2L3 as the most compelling candidate consistently ranked in the top 10 by three models, including state-of-the-art (SOTA) generalist models (gemma4-e4b, gemma4-12b-unified) and a model fine-tuned for drug target discovery (txgemma-9b-chat) [9].

**Table 2.**
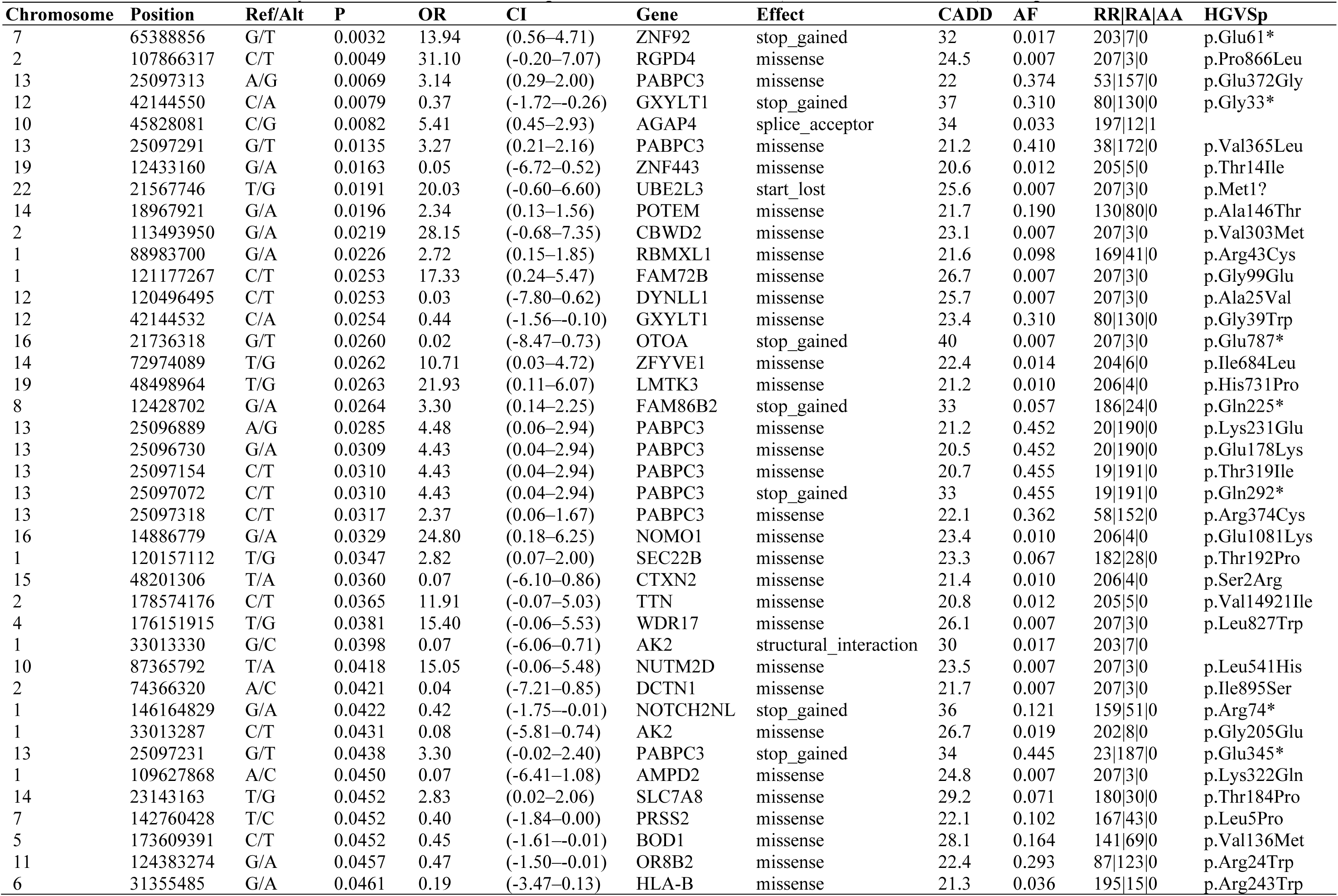
Forty variants from GWAS Unique to Qatari ICU Cohort (Absent from All of Us) with p<0.05 and CADD >20.

The associated and predicted deleterious founder allele at UBE2L3 (chr22:21,567,746:T>G; c.2T>G; p.Met1?) was identified with the following properties: CADD score of 25.6 (top 0.3% most deleterious), SIFT score of 0.001 (deleterious), allele frequency of 0.0071, allele count of 3, all heterozygous. Single variant and gene burden test results was OR=20.03, p=0.02, 95% CI = [.0.56, 718]. The variant affects the second nucleotide of the start codon (ATG → AGG), converting the initiator methionine to arginine and predicted to abolish translation initiation, resulting in complete loss of function of the 152-amino acid UBE2L3 protein, which contains a ubiquitin-conjugating (UBC) domain with an active-site cysteine residue.

All three heterozygous carriers were identified among sepsis cases and none among control (non sepsis ICU) (3/68 *vs* 0/142; Fisher’s exact p=0.026), though this observation is based on a small number of carriers and should be interpreted cautiously pending replication in larger cohorts. All carriers were male. The variant was absent from more than 945,000 individuals in gnomAD and the AoU database, consistent with an Arab-specific founder allele. Read-level verification using IGV confirmed the variant in all three carriers with adequate sequencing depth (Pgx-074: 22×, Pgx-087: 19×, Pgx-167: 40×), with alternate allele reads visible on both forward and reverse strands (Supplemental Figure 3).

### Independent Gene-Level Validation of *UBE2L3*

Because variant-level replication was not possible for this apparently population-specific allele, the gene was evaluated independently in the AoU. Gene-based burden analysis of UBE2L3 missense variants in AoU participants with bacterial infection (N=26,939 cases; N=243,470 controls) demonstrated a significant association (SKAT-O P = 2.38×10⁻³, OR=23.10, 95% CI= [0.63,845.27]), providing independent gene-level genetic support for a role of UBE2L3 variation in infection susceptibility.

### Overlap with Published Sepsis GWAS

Comparison with the GWAS Catalog, which contained 62 genes associated with sepsis-related traits (Supplemental Table V, VI), identified 25 genes from our study results that overlapped with previously reported (Supplemental Table VII) associations. These included *USP37, LRBA, KIF15, GPM6A, MAD1L1, RIN3, NFKBIL1, CHRNA7, SLAMF6, LINC00378, SAMD9, SLC35F4, TFRC, CLIC6, ANKMY2, CSMD1, FER, RPE65, HRH1, LPP, LINC00887, GAK, VPS13A, CRISPLD2* and *FLT1*.

### Cross-Referencing with All of Us

Among the 316 variants with CADD ≥20 and p ≤0.05 in this study, 17 variants showed associations in AoU (Supplemental Table VIII, IX). Additionally, 117 variants had homozygous carriers in the HMC ICU cohort. Twelve variants showed female-specific significance (p<0.05 in females, p>0.05 in males), and 58 variants across 57 genes were common in the ICU cohort but rare in gnomAD, suggesting population-specific enrichment.

Burden testing of the 295 genes with CADD ≥20 and p ≤0.05 variants from GWAS identified 163 genes with burden test p<0.05 (Supplemental Table X). HLA-DQA1[33] and PSMB8[34] showed the strongest biological connections to sepsis pathophysiology. ABCG2 has a mechanistic connection to antibacterial immunity and macrophage biology relevant to sepsis [35]. SBNO1 has limited direct functional evidence in sepsis; however, proteomic profiling has identified SBNO1 among proteins associated with sepsis-specific plasma peptide signatures, supporting its potential involvement in the systemic host response to severe infection [36]. Among the 163 genes with p<0.05 there were 12 genes containing 32 variants unique to the ICU cohort: *FAM72B, NOTCH2NL, SEC22B, BOD1, HLA-B, ZNF92, DYNLL1, GXYLT1, PABPC3, ZFYVE1, ZNF443*, and *UBE2L3*.

### Regional Linkage Disequilibrium Analysis of the *UBE2L3* Locus

To determine whether the observed association at UBE2L3 (chr22:21,567,746 T>G; c.2T>G; p.Met1?) could be attributed to linkage disequilibrium (LD) with a nearby common variant rather than the rare start-loss allele itself, we performed regional LD analysis across a 350-kilobase window (chr22:21,400,000–21,750,000; GRCh38) centered on the index variant (Table 3). Phased r² was computed against the index variant for all 3,620 biallelic single-nucleotide variants within the region using phased WGS haplotypes from all 210 study participants (PLINK2 v2.0.0-a.7.1 --r2-phased; Figure 3A).

**Figure 3.**
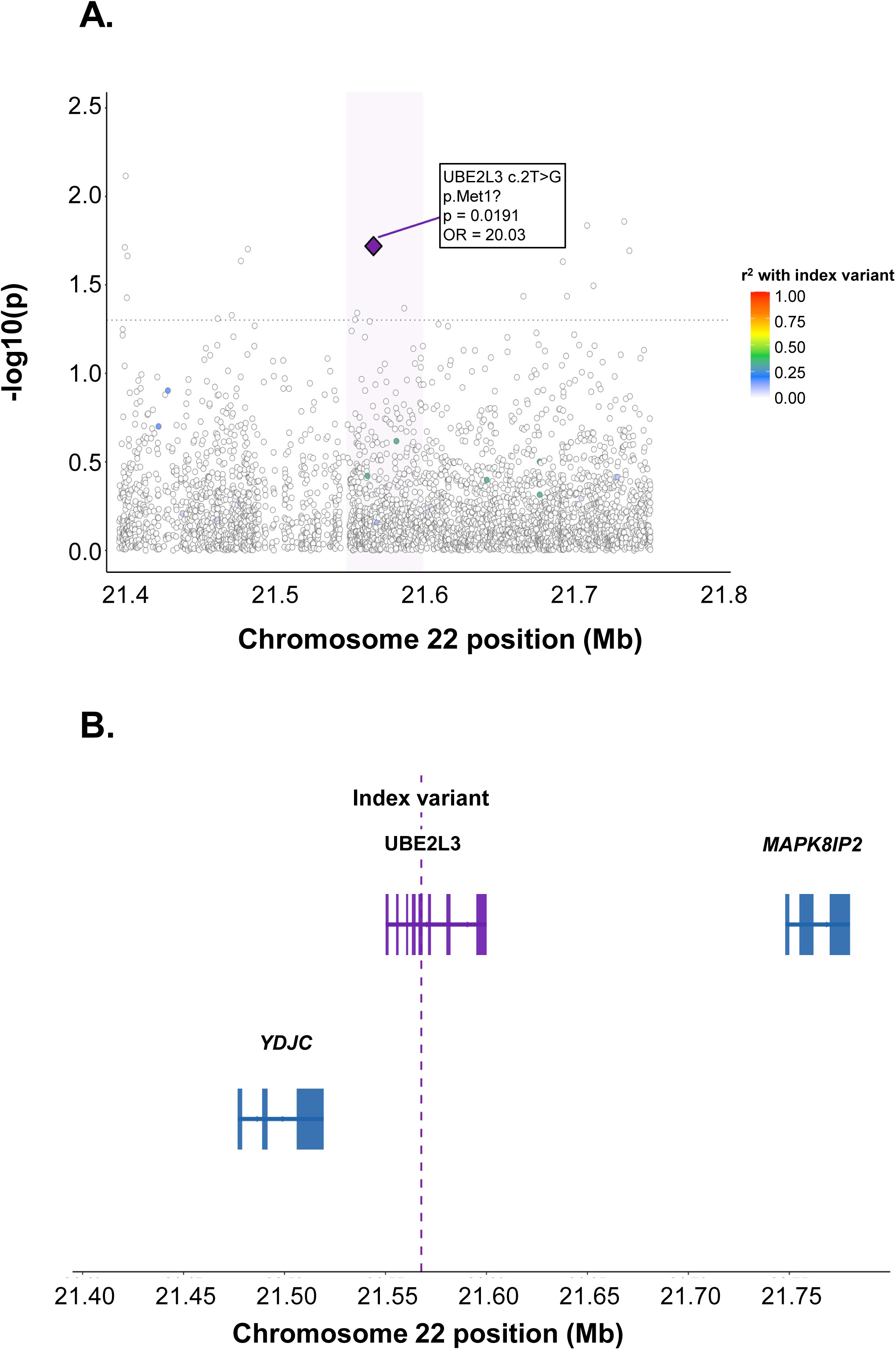
Regional association and linkage disequilibrium plot of the ***UBE2L3*** locus. **A.**-log₁₀(P) for 3,620 biallelic SNPs within the chr22:21.4–21.76 Mb region (GRCh38). Each point represents one variant; color indicates phased r² with the index variant (chr22:21,567,746 T>G; purple diamond) computed from WGS haplotypes in 210 Arab ICU patients (PLINK2 v2.0.0 -- r2-phased). Purple shading indicates the *UBE2L3* gene body. Dotted horizontal line indicates nominal significance (P=0.05). The absence of variants in strong LD (maximum r²=0.332; no variant with r²>0.5) confirms that the association signal is driven by the rare start-loss allele rather than a linked common variant, consistent with an Arab-specific founder allele on a private haplotype. **B.** Gene structure at the *UBE2L3* locus (Ensembl GRCh38). Exons shown as filled rectangles; introns as horizontal lines with directional arrows indicating transcriptional orientation. Purple dashed vertical line marks the index variant position. GRCh38/hg38 coordinates.

**Table 3.** All Variants with Phased r²>0.1 with the Index UBE2L3 Variant (chr22:21,567,746 T>G; p.Met1?) within the 350 kb Regional Window^1^.

| **Position (GRCh38)** | **$r^2$ (phased)** | **Nearest gene** | **Functional annotation** |
| --- | --- | --- | --- |
| chr22:21,563,817 | 0.332 | UBE2L3 | Intronic |
| chr22:21,582,819 | 0.332 | UBE2L3 | Intronic |
| chr22:21,642,326 | 0.332 | BCR | Intergenic |
| chr22:21,677,073 | 0.332 | BCR | Intronic |
| chr22:21,677,089 | 0.332 | BCR | Intronic |
| chr22:21,426,321 | 0.164 | YDJC | Intergenic |
| chr22:21,432,472 | 0.164 | YDJC | Intergenic |
| chr22:21,614,219 | 0.164 | BCR | Intronic |
| chr22:21,570,124 | 0.108 | UBE2L3 | Intronic |

<sup>1</sup> Variants are sorted by $r^2$ in descending order. No variant reached $r^2 > 0.5$ .

The LD structure at the UBE2L3 locus was notably sparse. The maximum observed r² between the index variant and any other variant in the 350 kb window was 0.332 (chr22:21,563,817 C>T; chr22:21,582,819 C>T; chr22:21,642,326 G>A; and chr22:21,677,073 G>A; all r²=0.332), and only nine variants across the entire region exceeded r²>0.1 with the index variant (Table 3). No variant reached r²>0.5, and the vast majority of the 3,620 variants tested showed r²<0.01 (Figure 3A). The four variants with maximum r² (r²=0.332) were intronic or intergenic variants with no predicted functional consequence and are most parsimoniously explained by shared haplotype background among the three heterozygous carriers rather than independent association signals.

The absence of variants in strong LD (r²>0.5) with the index variant has two important biological implications. First, it confirms that the UBE2L3 p.Met1? association signal cannot be explained by a common variant in LD; the signal is driven by the rare start-loss allele itself. This is consistent with the ultra-rare allele frequency of the variant (AF=0.007 in our cohort; absent from gnomAD and the AoU), which precludes the accumulation of strong LD with common variants across generations. Second, the sparse LD architecture is characteristic of a founder allele on a private haplotype, a variant that entered the population through a single founding event and has not had sufficient time or population size to accumulate LD with surrounding common variants at the population level. The low background r² across the locus (median r²<0.001 for the 3,620 variants tested) further supports the interpretation that the three heterozygous carriers share a recent common ancestor on the Arab founder haplotype.

The regional gene structure (Figure 3B) shows that the index variant falls within the first exon of UBE2L3 at the translation start site, flanked upstream by YDJC (chr22:21,476,842– 21,519,400; C22orf39; ubiquitin-binding domain-containing protein) and downstream by the breakpoint cluster region BCR (chr22:21,620,000+). Neither flanking gene contains variants in strong LD with the index variant and no YDJC or BCR variants showed functional annotations (CADD≥20) at positions in LD with the index variant.

Taken together, the regional LD analysis establishes the UBE2L3 p.Met1? start-loss variant as the credible causal variant at this locus. The 95% credible set for this association contains a single variant the index variant itself with no common variant proxy capable of mediating the observed association. This fine-mapping resolution, achieved without formal Bayesian fine-mapping due to the low LD background, is a direct consequence of the rare founder allele architecture and represents a significant analytical advantage of studying disease susceptibility in Arab founder populations: rare deleterious variants that have not been diluted by admixture can be localized to single-variant resolution, providing unambiguous targets for functional characterization.

## Discussion

In this genome-wide association study of sepsis susceptibility using WGS in 210 Arab ICU patients, we identified a compelling Arab founder loss-of-function variant in UBE2L3 (c.2T>G; p.Met1?) with independent gene-level validation. This variant was absent from 14,669 individuals in the Qatar Genome Programme [8] and from gnomAD v4.1 (251,000 individuals), confirming its status as an Arab-specific founder allele absent from reference databases and invisible to conventional GWAS conducted in European-ancestry populations.

UBE2L3 encodes an E2 ubiquitin-conjugating enzyme that plays a critical role in the ubiquitination of the NF-κ B precursor p105, placing it directly in the TLR→NF-κ B→cytokine signaling axis that is central to the innate immune response to infection [37]. The start-loss variant is predicted to abolish translation initiation, resulting in complete loss of function. UBE2L3 has been associated with several autoimmune diseases through GWAS, including rheumatoid arthritis, celiac disease, Crohn’s disease and systemic lupus erythematosus, and has been linked to natural killer cell cytotoxic function and clearance of chronic hepatitis B virus infection [38]. While it is known that sepsis is a leading cause of morbidity and mortality among autoimmune disease patients, the biological mechanisms behind that relationship are poorly understood [39].

The potential relevance of UBE2L3 to sepsis is particularly supported by its role in NF-κB signaling, a central pathway in the host response to infection. UBE2L3 has been identified as a functional E2 partner of the linear ubiquitin chain assembly complex (LUBAC), where it promotes linear ubiquitination of NEMO and facilitates recruitment and activation of the NF-κB signaling complex following TNF-α stimulation. Experimental depletion of UBE2L3 reduces NEMO linear ubiquitination, NF-κB activation, and downstream inflammatory gene expression, demonstrating that UBE2L3 can directly modulate cytokine-driven inflammatory signaling. Because dysregulated NF-κB activation contributes to the excessive inflammatory response characteristic of sepsis, alterations in UBE2L3 function could plausibly influence the magnitude or persistence of host inflammatory responses during systemic infection [38, 40].

UBE2L3 also regulates IL-1β by targeting it for degradation, providing an additional mechanistic link between UBE2L3 depletion and sepsis. Studies in human and murine macrophages have shown that UBE2L3 promotes K48-linked ubiquitination and proteasomal turnover of pro-IL-1β, thereby limiting the amount of substrate available for caspase-1-mediated processing. Conversely, depletion or caspase-1-mediated loss of UBE2L3 increases pro-IL-1β abundance and enhances secretion of mature IL-1β following inflammasome activation. This function has been demonstrated in response to bacterial infection, including both Gram-negative and Gram-positive organisms. These observations suggest that altered UBE2L3 activity could affect the balance between protective antimicrobial inflammation and excessive cytokine production, a critical determinant of sepsis pathophysiology [10].

The convergence of multiple lines of evidence supports UBE2L3 as a sepsis susceptibility gene: (1) predicted complete loss of function due to start-codon disruption; (2) high deleteriousness scores (CADD = 25.6, SIFT = 0.001); (3) exclusive presence in sepsis cases (3/68 *vs* 0/142 controls; Fisher’s exact pvalue = 0.026; noting that ICU-based case-control ascertainment introduces selection bias and the carrier frequency cannot be extrapolated to the general Arab population); (4) absence from >945,000 individuals in gnomAD and AoU; (5) established biological role in NF-κB signaling and innate immunity; (6) independent gene-level validation in AoU (SKAT-O P = 2.38×10⁻³); and (7) top-ranked drug target by an LLM fine-tuned for pharmaceutical target attractiveness.

### Limitation

This study has several limitations. First, the modest sample size of 210 patients limited statistical power, and no variants reached genome-wide significance. The suggestive associations require replication in larger cohorts. Second, the UBE2L3 finding, while biologically compelling, is based on only three carriers and requires functional validation. Third, the study cohort was drawn from a single ICU in Qatar, and the generalizability of findings to other populations is uncertain. Fourth, the case-control imbalance (32% cases) may reduce power despite the use of Firth logistic regression to address this. Fifth Case-control ascertainment within a single tertiary ICU introduces selection bias, as controls represent critically ill non-sepsis patients rather than healthy population controls, limiting generalizability of carrier frequency estimates to the broader Arab population. Finally, the use of LLM-based target prioritization is exploratory and should not be interpreted as definitive validation.

Despite these limitations, this study demonstrates the power of WGS in founder populations to identify rare, high-impact variants that would be missed by conventional genotyping approaches. The identification of population-specific variants with large effect sizes is particularly relevant for precision medicine approaches to sepsis in diverse populations.

## Supporting information

Supplemental figures, legends and list of tables

## Acknowledgments

We thank all Qatar Research ICU Network collaborators: Abdelraouf Akkari, Awab El Shaikh, Ans Alamami, Abdulaziz Al-Alawi, Mohammed Qandil, Rabee Tawel, Ahmad Al Johari, Mohammed Faris, Anoud Duale, Khaled Ghazwi, Mojahid, Abdurrahman El Buzidi, Awadh Bin Taher, Ezzeddin Ibrahim and Nasseem Albadw for helping with patient recruitment, and N. Mohamed for editorial support. These studies were supported, in part, by Qatar Research Development and Innovation Council PPM 03-0314-190024 and Department of Genetic Medicine, Weill Cornell Medicine. The Qatar Biobank is funded by the Qatar Foundation for Education, Science and Community Development and the Supreme Council of Health.

AI-based tools were used solely to assist with grammar, spelling, and language editing. The authors reviewed and approved all edited content and take full responsibility for the final manuscript.

## Competing interests

None.

## Author contributions

Conceptualization: JRF, MRR; Data curation: AR, AAH; Formal Analysis: JRF, MRR, JGM; Funding acquisition: RGC, AR; Investigation: MRR, JRF, AR; Methodology: MRR, JRF; Project administration: AR, RGC; Resources: AAH; Supervision: ES, RGC; Validation: JGM; Writing - original draft: MRR, JRF; Writing – review & editing: RGC

## Data availability

All data produced in the present study are available upon reasonable request to the author

## Notes

### Competing Interest Statement

The authors have declared no competing interest.

### Author Declarations

The Ethics committee (Internal Review Board) of Hamad Medical Corporation (HMC) gave ethical approval for this work (IRB 19-00037).

