## Supplemental figures, legends and list of tables for "Genome-Wide Association Study of Sepsis Susceptibility in ICU Patients Identifies Arab Founder Loss-of-Function Variant in *UBE2L3*"

#### Supplemental Figure Legends

**Supplemental Figure 1.** QQ plot demonstrating genomic inflation control. Observed *vs* expected -log10(p-values) for 16.6 million variants. The genomic inflation factor $\lambda\_GC = 0.840$ indicates excellent control of population stratification with no systematic bias.

**Supplemental Figure 2.** QQ plot demonstrating calibration of gene-level burden test statistics. Observed *vs* expected -log10(P-values) for 20,412 genes. The genomic inflation factor ($\lambda\_GC = 0.789$) indicates modest deflation of the test statistics, suggesting conservative calibration rather than systematic genomic inflation.

**Supplemental Figure 3**. IGV read-level verification of the candidate variant in three carriers. Integrative Genomics Viewer (IGV) screenshots confirm the presence of the variant in all three with adequate sequencing depth (22×, 19×, and 40×, respectively). Alternate allele reads are observed on both forward and reverse strands, supporting the authenticity of the variant and reducing the likelihood of strand-specific sequencing artifacts.

**Supplemental Figure 1**

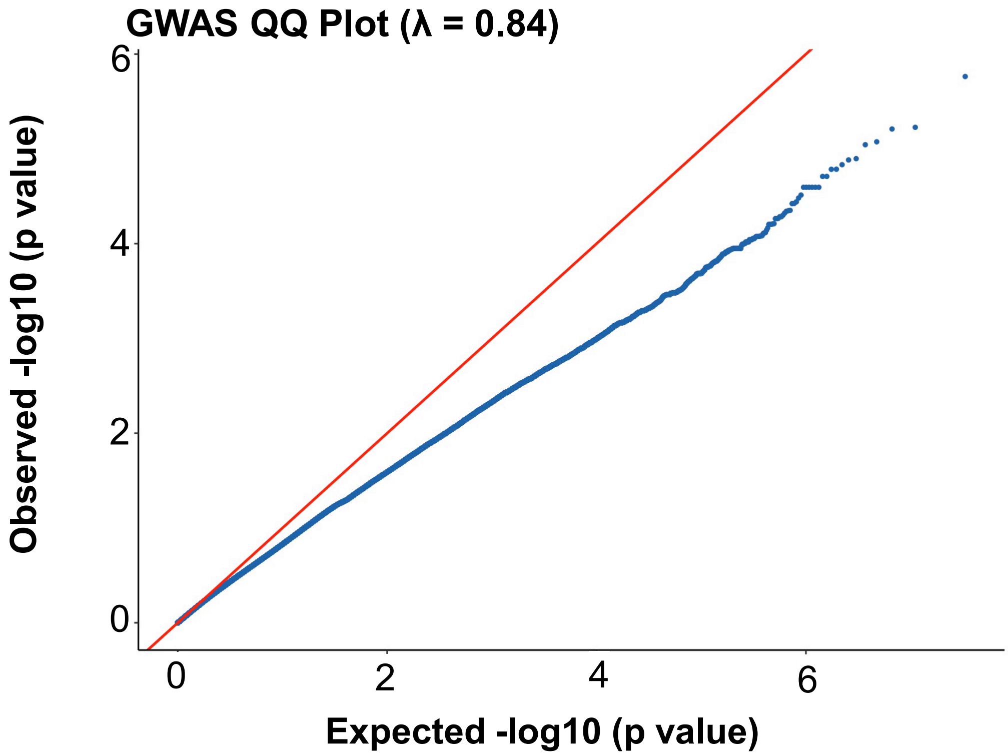

**Supplemental Figure 2**

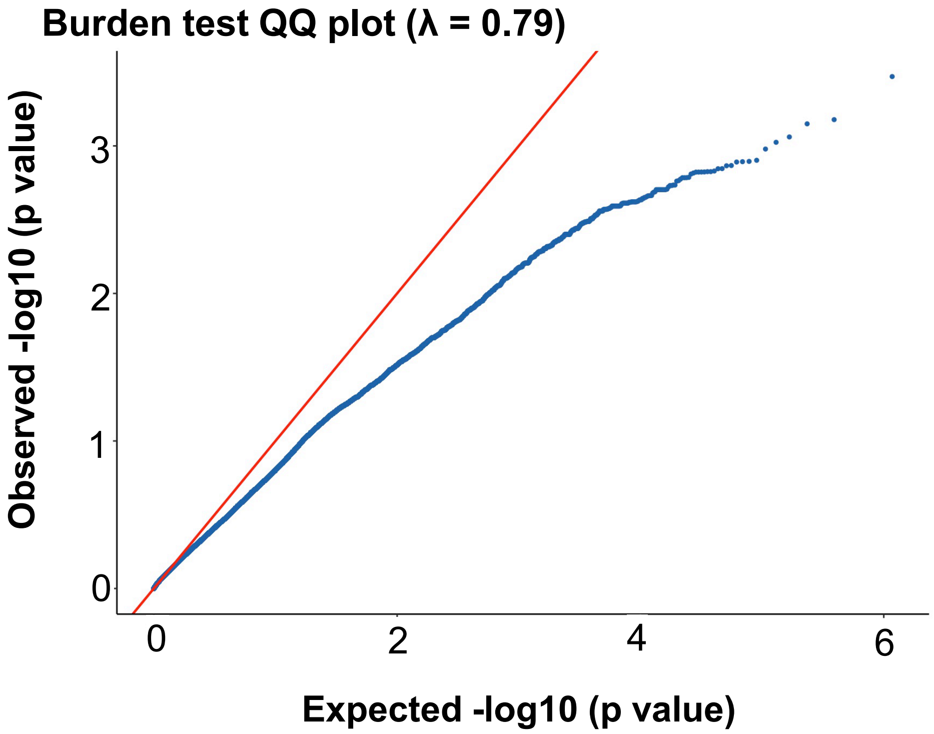

### Variant position at the first codon of UBE2L3

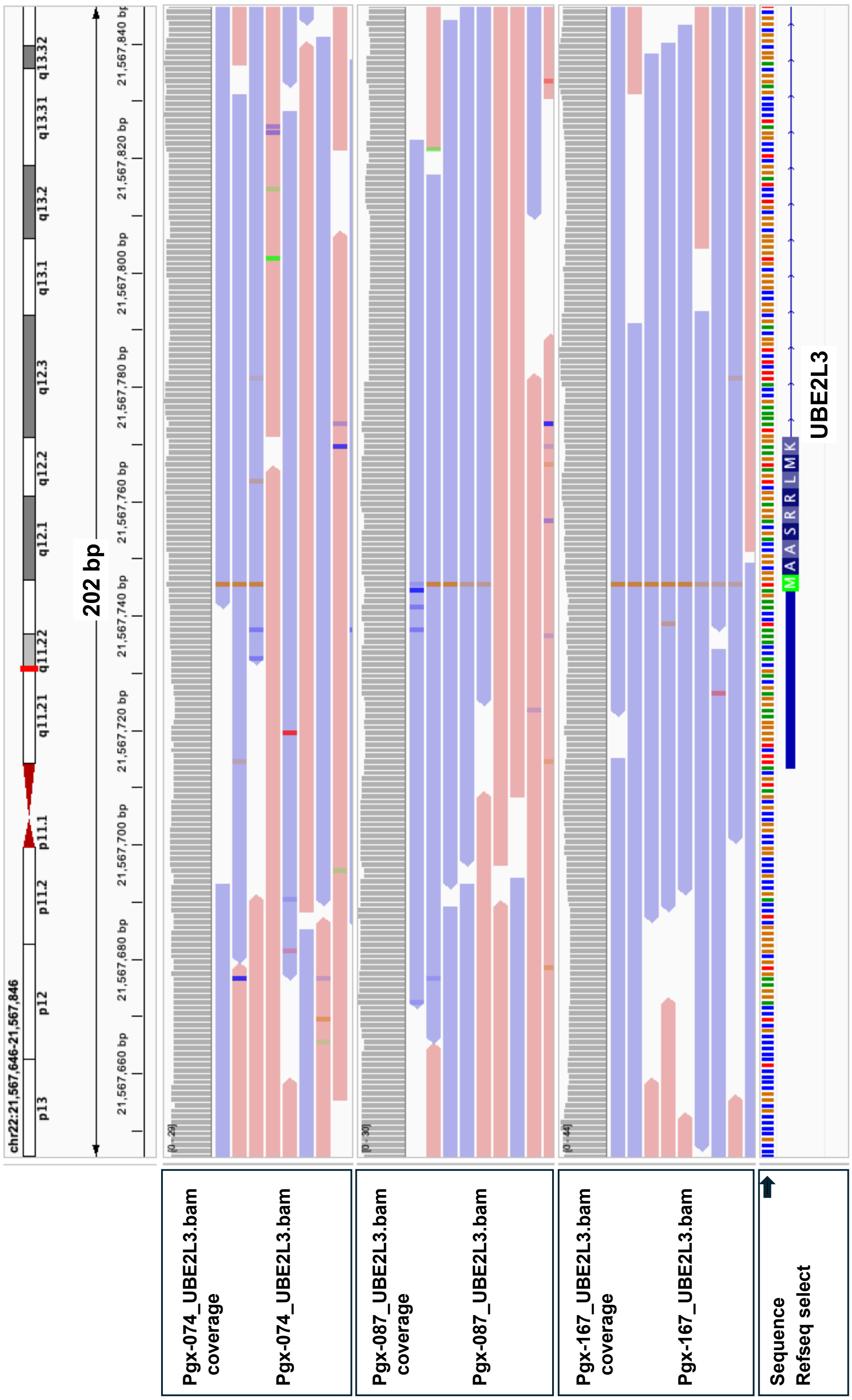

#### Supplementary Material

| | |
|---|---|
| **Supplementary Table I.** | 316 variants from GWAS with p≤0.05and CADD ≥20 |
| **Supplementary Table II.** | 295 gene summaries for variants from GWAS with CADD ≥20 and p ≤0.05 |
| **Supplementary Table III.** | Forty 0 variants from GWAS with p≤0.05 and CADD ≥20 unique to Qatari ICU Cohort (Absent from All of Us) |
| **Supplementary Table IV.** | Burden test analysis for 40 variants from GWAS unique to Qatari ICU association with genes with p ≤0.05 Supplementary Table V. GWAS catalog downloaded from GWAS catalog "gwas-association-downloaded_2026-07-23-HP_0100806-withChildTraits.tsv" |
| **Supplementary Table VI.** | GWAS catalog summary |
| **Supplemental Tabe VII.** | 25 Genes found form GWAS catalog in Qatari ICU cohort |
| **Supplemental Table VIII.** | 17 variants from GWAS analysis with p≤0.05and CADD ≥20 found in AoU GWAS association |
| **Supplementary Table XI.** | 17 variants from GWAS analysis with p≤0.05and CADD ≥20 found in AoU GWAS association |
| **Supplemental Table X.** | 163 genes with burden test p≤0.05 |
